# A mixed methods study protocol to develop and pilot a Competency Assessment Tool to support therapists in the care of patients with blunt CHest trauma (CATCh Study)

**DOI:** 10.1101/2025.07.10.25325601

**Authors:** Ceri Battle, Jane Barnett, Timothy Driscoll, Hayley Hutchings, Claire O’Neill, Hannah Toghill, Rhys Whelan, Edward Baker

## Abstract

**Introduction:** Competency Assessment Tools are well-recognised as a method to achieve a standardised level of practice for a group of healthcare professionals with similar characteristics. The aim of this study is to develop and pilot a new Competency Assessment Tool to support therapists caring for patients with blunt chest trauma from pre-hospital care through to long-term follow-up following hospital discharge.

**Methods and analysis:** A mixed methods study will be undertaken, with three distinct phases; 1) an integrative narrative review to examine the literature regarding therapist competencies (described elsewhere), 2) focus groups with patients, therapists and key stakeholders to explore opinions regarding important aspects of care (phases one and two will inform the content of the tool), followed by final tool development by an international expert panel, and 3) a multi-centre pilot study using questionnaires and elicitation interviews, in which final tool acceptability to therapists will be tested. The total sample size will be between 40-50 participants for the focus groups. For the final tool development work, a panel of 10 international experts will be identified, with a subgroup of three to five experts who will be recruited to confirm content validity. We will pilot the tool at five health boards in Wales, aiming for 10 therapists from each. Elicitation interviews will be undertaken with a smaller sample size of between 15-20 therapists. A mixed qualitative and quantitative data analysis approach will be used.

**Ethics and dissemination:** Proportionate ethics approval has been granted (Reference number: 24/YH/0231). We will publish the work in an open access peer reviewed journal to ensure equitable access and present at relevant conferences. Webinars will be used to achieve a wide audience. The results will be shared with the research participants via an infographic which will be designed and developed with the public research partners.

**Study registration:** Integrative review is registered at the Open Science Framework: https://doi.org/10.17605/OSF.IO/CEXNR

**STRENGTHS AND LIMITATIONS OF THIS STUDY:** Harnessing the expertise of patients, therapists and stakeholders, this mixed methods study will develop and pilot a Competency Assessment Tool for therapists managing adults with blunt chest trauma.

The use of questionnaires and elicitation interviews in the pilot study will facilitate an indepth exploration of therapist acceptability of the new tool.

The new tool will potentially lead to improved standardisation of care for patients with blunt chest trauma.

The integrative narrative review will potentially be limited by a lack of published research focussed on therapy for patients with blunt chest trauma.

Ensuring content validity of the tool could be challenging, due to the wide inclusion criteria of the proposed tool.

## INTRODUCTION

## Background

Difficulties in the overall management of patients with blunt chest trauma are very well-reported, from the pre-hospital care setting through to long-term follow-up months and even years after hospital discharge.[1] Patients with blunt chest trauma range between a young fit adult following a road traffic incident, to an older adult with frailty who has sustained a fall from a standing height. Therapists provide a range of interventions for patients with blunt chest trauma, broadly categorised into respiratory care and rehabilitation.[2] In the early stages post-injury, using a variety of techniques and devices the therapists will be responsible for assisting the patient with maintenance of adequate lung expansion and clearance of pulmonary secretions. In this early stage, rehabilitation will target safe and early mobilisation, and restoration of normal posture and movement patterns. In the later stages of recovery, respiratory care will focus more on patient education and promotion of self-management. Rehabilitation will include upper limb strengthening, general conditioning and a return to full baseline functional status.[2,3] This management will also vary according to the patient’s presentation.

A global survey reported that patients with major chest trauma commonly appear to be treated by physiotherapists with breathing exercises and mobilisation activities; it is also noted that post-discharge rehabilitation is rare despite evidence that these patients experience ongoing pain and physical disability.[3] It was concluded that limited research exists regarding the role of therapy in the management of patients with blunt chest trauma and consequently, there is a lack of standardisation in care.[3] In a recent international e-Delphi study to develop guidance for physiotherapists managing patients with blunt chest trauma, a key conclusions was that further research is needed to standardise therapy management for these patients, and subsequently to identify training needs for therapists managing this complex group of patients.[4]

Competency Assessment Tools (CATs) are well-recognised as a method to achieve a standardised level of practice for a group of healthcare professionals with similar characteristics.[5-7] Competencies can be described as a combination of observable and measurable knowledge, skills, abilities, and personal attributes that make up a clinician’s performance.[8] In the healthcare system, core competency standards are the criteria and requirements used to inform standards of practice of the specific profession.[8] It is the responsibility of each clinician to be competent in delivering the skills required to improve and sustain the quality of patient care. Evidence also highlights the importance of healthcare organisations assessing the clinicians’ professional competencies. There is a need therefore for valid and reliable assessment tools. Currently, no such tool exists for therapists managing patients with blunt chest trauma, and as a result, a lack of standardisation in care exists in the UK.[9]

### Study aims

The aim of this study is to develop and pilot a new CAT to support therapists caring for patients with blunt chest trauma from pre-hospital care (care received in the community before arriving at the hospital) through to long-term follow-up (care received in the community following hospital discharge). In order to achieve this aim, the following objectives will be addressed:

1. Complete an integrative review of the literature describing the role of the therapist in the management of patients with blunt chest trauma (published elsewhere).
2. Undertake focus groups with therapists, patients and key-stakeholders to explore potential domains and items for the CAT.
3. Develop the CAT using a nominal group technique with a panel of 10 international experts.
4. Pilot the CAT at five Welsh health boards using questionnaires and elicitation interviews.

## METHODS AND ANALYSIS

### Study design

A mixed methods study will be undertaken, with three distinct phases to collate both qualitative and quantitative data. These will include an integrative narrative review (published elsewhere), focus groups with therapists, patients and key stakeholders, and a multi-centre (at five health boards in Wales) pilot study of the CAT (including questionnaires and elicitation interviews) in which acceptability to therapists will be tested. The six-step model guidance for developing CATs has been used to develop this protocol.[6]

#### Participants

Adults with lived experience of being patients with blunt chest trauma, and all allied health professional therapists and clinical stakeholders who are involved in the direct care of these patients from pre-hospital care to long-term follow-up (defined as care received before arriving at hospital, during any hospital attendance and following hospital discharge). This will include therapists working in various care provision teams in the community or hospital setting and from different geographical locations within Wales, reflecting the diversity and underserved groups in society. Using the INCLUDE Framework as a guide[10], representation and inclusivity will be ensured, in terms of gender, disability, race, religion, social status, neuro-diversity and sexuality.

#### Inclusion criteria

- Currently employed as a therapist (Allied Health Professional) working with patients with blunt chest trauma, working in either primary, secondary or tertiary care setting.
- Stakeholders: clinical professionals who are responsible for the care of patients after chest wall injuries these may include (but not limited to): Trauma Surgeons, Trauma/Emergency Care/Critical Care physicians, Geriatricians, Advanced Clinical Practitioners, Pharmacists, Registered Nurses, Paramedics, General Practitioners
- A member of the public who has been a patient with blunt chest trauma requiring hospital care, in the last three years (regardless of whether admission to hospital was required or not).

#### Exclusion criteria

- Healthcare professionals not involved with the care of patients with blunt chest trauma.
- Therapy students (not yet qualified).
- Patients lacking sufficient capacity to participate in the focus group (without support of a family member / carer).

#### Setting and recruitment

Phase 1: Not applicable as this phase will involve completing an integrative review.

Phase 2: Participants (therapists, public research partners and key stakeholders).

CB is currently completing a coproduction project, in which a large and diverse group of public research partners (adults with lived experience of being patients with blunt chest trauma) are engaged. This group will be approached to consider participating in the phase two focus groups. Additional patients will be approached and asked to consider participating in the focus groups while under the care of the physiotherapy chest trauma team conducting this study. They will be provided with a Participant Information Sheet at the point of recruitment, with consent to participate in the focus group taken at the focus group meeting, either verbally if online (and signed by the researcher facilitating the meeting), or in writing if the meeting is face to face. Phase 2 also involves recruiting therapists and key stakeholders. Adverts and clinician participant information sheets will be sent to the therapy teams managing the patients with blunt chest trauma in five Welsh participating Health Boards.

Consent will be sought and obtained verbally and recorded at the start of each focus group meeting.

Phase 3: Participants (therapists piloting the CAT).

Participants will be recruited by local principal investigators (PIs) at each of the five participating Health Boards. A Clinician Participant Information Sheet will be provided and the study discussed with the site’s Principal Investigator (PI). Written consent to complete the CAT and feedback survey will be obtained by the PI from the therapist participants. At the point of completion of the tool and feedback survey, a small sample of therapists from each hospital will be invited by the PI to participate in an online elicitation interview (approximately one hour) and provided with a clinician information sheet outlining study processes for the interview. Consent will be sought and obtained verbally and recorded at the start of each interview.

### Sample size

Phase one: Not applicable as this phase will involve completing an integrative review.

Phase two: Based on methodological guidance, it is proposed that the total sample size will be between 40-50 participants for the focus groups. In relation to competency development there is no defined ‘gold standard’ for sample size, therefore, the proposed sample was based on previous competency development studies.[5,7] For the final tool development work, a panel of 10 international experts will be identified, with a subgroup of three to five experts who will be recruited to confirm content validity.[11]

Phase three: We will pilot the tool at five health boards in Wales, aiming for 10 therapists from each. This will give an overall sample size of 50 therapists. Methodological guidance recommends that as the pilot study is not conducted for the purpose of inference, a power calculation is not needed.[12,13] The sample size does, however, need to be justified on the basis of the goal of the pilot study which, in this instance, is to test that the competencies included in the tool are expressed in a manner that is easily understood, recognizable, and demonstrable in professional practice.[14] The survey responses completed by the sample will allow us to evaluate clinician perceptions of the tool itself, whilst taking into account our pre-determined sampling framework, aiming for a diverse sample from across Wales. Elicitation interviews will be undertaken with a smaller sample size of between 15-20 therapists.

### Interventions

Phase one: published elsewhere

Phase two: Focus group meetings will be undertaken in which the participants will be asked to consider the key aspects of patient care, including knowledge and skills they feel are relevant to managing the patients with blunt chest trauma. Patients will be asked to consider the important aspects of their own blunt chest trauma experience and therapy received (if any).

The development of the tool will then be undertaken using a nominal group technique, following the guidance outlined in Harb et al (2021).[17] In the first instance, the research team will collate a list of domains and items, based on evidence gained in the review and focus groups. A panel of 10 international experts will meet via MS Teams, to discuss and vote on the proposed domains and items. The experts will eliminate items that are agreed not to be important, thus generating the shortlist of domains/items for the final tool. Content validity of the final tool will be confirmed using a subgroup of experts who, in a second MS Teams meeting, will review and rate for relevancy, readability and clarity. We will use a method described by Polit et al (2007), in which item-level content validity indexes will be translated into values of a modified kappa statistic.[11]

Phase three: The tool will then be piloted at five health boards in Wales, where questionnaires will be distributed to a diverse convenience sample of therapists working in a) prehospital care teams (such as those based in GP practices or paramedic teams), b) hospital settings (such as emergency care, frailty and ortho-geriatric services, respiratory and cardiothoracic specialists) and c) community setting (such as rehabilitation teams, outreach or follow-up services). Elicitation interviews will also be conducted.

Figure 1 illustrates the study phases.

**Figure 1:**
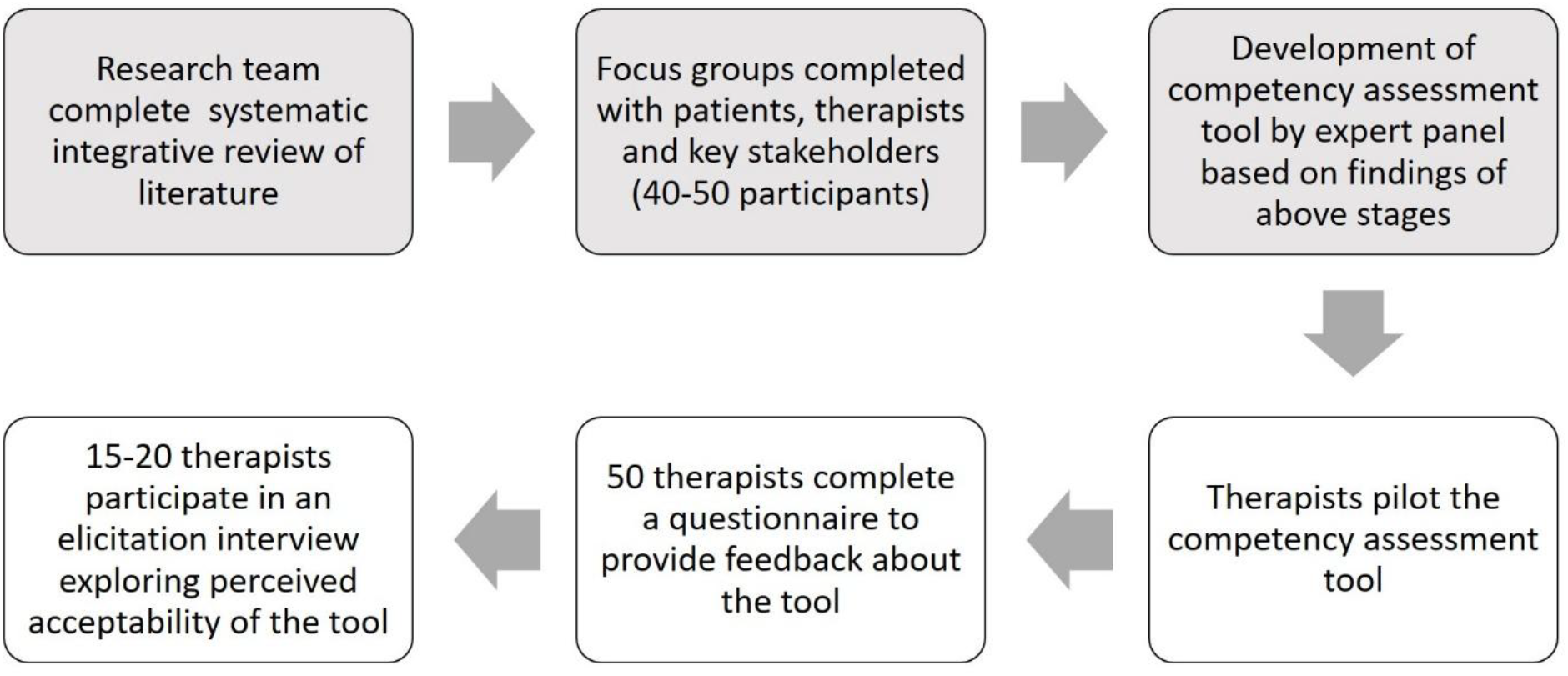
Study phases.

#### Data Management

Quantitative data will be managed using REDCap electronic data capture tools hosted at Swansea University.[18] REDCap (Research Electronic Data Capture) is a secure, web-based software platform designed to support data capture for research studies, providing 1) an intuitive interface for validated data capture; 2) audit trails for tracking data manipulation and export procedures; 3) automated export procedures for seamless data downloads to common statistical packages; and 4) procedures for data integration and interoperability with external sources. A database specification and data management plan will be agreed before recruitment and data collection commence.

Qualitative data will be managed using NVivo12 (computer assisted qualitative data analysis software).[19] Focus groups will be recorded with audio equipment if face to face or through MS Teams and transcribed in real time using the auto-transcribing function. This will be checked for accuracy by two researchers with the patient focus groups also being checked by a public research partner. Data from these focus groups will be anonymised prior to data analysis. Transcribed focus group data will be uploaded to NVivo12 in preparation for analysis. Public research partners will work with the qualitative research experts in the team by assisting with running the focus groups and commenting on code books and summaries as they are produced

#### Outcome measures

Phase two of this study will identify a template of key themes (rather than ‘outcomes’), which will form the basis of the CAT. The intervention development phase will be based on the Medical Research Council (MRC) framework.[20] In Phase three, the tool will be piloted on therapists working with patients with blunt chest trauma. Therapists will be asked to complete the CAT according to their current perceived level of competence, then complete a pre-designed and piloted questionnaire regarding their perceptions of the usefulness of the CAT. Elicitation interviews will also be conducted which will explore the participants’ attitudes towards the tool, appropriateness, suitability, convenience and perceived effectiveness of the CAT.

#### Data Analysis

Phase one: published elsewhere

Phase two: Qualitative data analysis will be undertaken by both the qualitative experts and public research partners in the team using Braun and Clarke’s process of reflexive thematic coding.[5,6] This method of analysis primarily identifies, examines and records data patterns from multiple similar data sets. These patterns are key in the description of the participants’ experiences, ideas, and opinions. Themes are identified by combining these data from individual interviews to build a comprehensive image of the collective experiences of the participants.[21,22] The analysis plan will broadly follow previously completed qualitative work on chest wall injury.[23] Transcribed interviews will be uploaded on the data management system NVivo v12 (QSR International Ltd) and initial data coding will be undertaken by two researchers.

Analysis will follow the follow six step process: 1) familiarisation with the data, 2) generating initial codes, 3) searching for themes, 4) reviewing themes, 5) defining and naming themes and 6) producing the report.[21] Consensus on codes and themes will be achieved through an integrated process of discussion between members of the research team. Trustworthiness and rigour are key to effective analysis using these methods. To ensure these values are maintained a trustworthiness criteria will be applied throughout the research process.[24] As the concept of data saturation is not recognised within reflexive thematic analysis, the evaluation of data saturation will not be undertaken, rather data collection and analysis will be pragmatically driven.[14,25]

Phase three: Quantitative analysis of the questionnaire Likert response data will be completed using McNemar’s test. Qualitative analysis of elicitation interviews will be completed as in phase three above, and again the public research partners will assist the qualitative experts with this process. The elicitation interviews will be used to capture the therapists’ narrative accounts of their engagement with the checklist. The interviews will be structured around the domains/fields of the checklist and the therapists will be asked to describe their own subjective interactions with the checklist in order to understand the meaning they attach to the process of completing it. The resulting data will be then analysed according to the principles of interpretative phenomenological analysis (IPA), which examines how a given person, in a given context, makes sense of a given situation.

IPA is rooted in phenomenology, and is based on a commitment to examining the detailed experience of each case, in turn, before making more general claims. It is particularly useful for looking at complex issues where the researcher and the participant are trying to make sense of their experience. The process involves multiple readings of the transcripts to create a descriptive account of the experience through the eyes of the participant.. At each stage themes are examined to highlight convergences and divergences to illuminate the breadth and depth of each. The next step is interpretation and the use of existing theories such as self-efficacy as determinants of behaviour and social cognitive theory that emphasises the influence of the social environment (including role modelling, re-enforcement and expectation) on behaviour. The researchers maintain a systematic and reflexive approach to the recognition of themes to ensure they can be traced back to the expressions of the research participants. A summary of themes and sub-themes will be reviewed by a non-participant therapist to confirm the analysis.[26] Quantitive and qualitative data will be integrated following a triangulation design and convergence model.[27]

#### Public Research Partner involvement

Two public research partners have been involved from the outset in this phase of work. They have advised us on aspects of the study design, including the inclusion of patients in phase two of the work, the choice of elicitation interview in phase three, and the diverse sampling framework throughout the study. Both representatives will be members of the study management group for the duration of the study. Experienced patient representatives will be involved in the data analysis process through the study and will be co-authoring outcomes to ensure that these remain inclusive and patient centred.

## Data Availability

All data produced in the present study are available upon reasonable request to the authors

## ETHICS AND DISSEMINATION

### Ethical issues

Proportionate ethics approval has been granted (Reference number: 24/YH/0231). Any necessary protocol modifications will be communicated to the investigators, regulatory authorities, study participants and study registries in a timely manner. Compliance with this will be monitored by the study sponsor Swansea Bay University Health Board (SBUHB) Research and Development Department. PIs will all be trained in Good Clinical Practice. Informed consent will be obtained by the research team or PIs who will all have received ‘protocol and informed consent specific training’ in alignment with the principles of GCP and who have signed the study delegation log.

Consent will be sought, following a full introduction to the study, prior to the start of the focus group. The study’s chief investigator will assume overall responsibility to ensure that participant anonymity is protected and maintained. Study data will be kept confidential and managed in accordance with the Data Protection Act, NHS Caldicott Guardian, The Research Governance Framework for Health and Social Care and Research Ethics Committee Approval. Once informed consent is obtained, all participants will be allocated a study pseudonym. The CI will act as the custodian of the data and the records will be kept securely for a further five years in the SBUHB archive facility. The Caldicott guidelines will be adhered to throughout phases two and three of the study.

### Dissemination policy

The principle output of this work will be the CAT itself, which we will publish in an open access peer reviewed journal (to ensure equitable access) and present at relevant conferences. The dissemination of the study results will facilitate the development of evidence-based knowledge and clinical skills necessary to deliver optimal patient care. Local, national and international seminars / webinars will be used to achieve a wide audience. The results will be shared with the research participants via an infographic (and wider on social media), which will be designed and developed by the research team with the public research partners. Any research participant who has expressed interest in the study findings will be sent a copy of the final infographic. We aim to disseminate the findings on an international scale, using the professional networks such as the Chartered Society of Physiotherapy and Royal College of Occupational Therapy, and the Chest Wall Injury Society (CWIS) Group for dissemination, which now has international representation.

## Authors contributions

CB and EB conceptualised the study. RW contributed to the design of the integrative review. JB contributed to the development of study design as a public research partner. All authors developed the study protocol. CB drafted the article and JB, TD, HH, CON, MP, HT, RW and EB all read, edited and approved the final version.

## Data statement

Data can be accessed from the corresponding author on reasonable request

